# Value of using artificial intelligence derived clusters by health and social care need in Primary Care: A qualitative interview study

**DOI:** 10.1101/2024.10.17.24315657

**Authors:** Sian Holt, Glenn Simpson, Miriam Santer, Hazel Everitt, Andrew Farmer, Kuangji Zhou, Zhiling Qian, Firoza Davies, Hajira Dambha-Miller, Leanne Morrison

## Abstract

**Purpose:** People living with MLTCs attending consultations in primary care frequently have unmet social care needs (SCNs), which can be challenging to identify and address. Artificial intelligence (AI) derived clusters could help to identify patients at risk of SCNs. Understanding the views of people living with MLTCs and those involved in their care can help inform the design of effective interventions informed by AI-derived clusters to address SCNs.

**Methods:** Qualitative study using semi-structured online and telephone interviews with 24 people living with MLTCs and 20 people involved in the care of MLTCs. Interviews were analysed using Reflexive Thematic Analysis.

**Results:** Primary care was viewed as an appropriate place to have conversations about SCNs. However, participants felt health care professionals lack capacity to have these conversations and to identify sources of support. AI was perceived as a tool that could potentially increase capacity for this but only when supplemented with effective, clinical conversations. Interventions harnessing AI should be brief, be easy to use and remain relevant over time, to ensure no additional burden on clinical capacity. Interventions must allow flexibility to be used by multidisciplinary teams within primary care, frame messages positively and facilitate conversations that remain patient centered.

**Conclusion:** Our findings suggest that AI-derived clusters to identify and support SCNs in primary care have perceived value, but there were some concerns including the need to consider personal context. AI derived clusters can be used as a tool to inform and prioritise effective clinical conversations.

**Conference names, dates and locations for any prior presentations:**

- British Journal of General Practice Research Conference, March 2024, London.
- School of Academic Primary Care Southwest Conference, March 2024. Cardiff.
- Faculty of Medicine Research Conference, June 2024. Southampton.

## Introduction

Multiple Long-Term Conditions (MLTCs) are defined as a person having two or more health conditions(1). MLTCs are costly for the NHS and associated with lower quality of life and worse mental health for patients (2, 3). People living with MLTCs often have complex or unmet social care needs (SCNs) that require support (4). Previous evidence shows lower rates of full-time employment and greater need for housing and financial assistance among people living with MLTCs (5). Integrating care to more holistically manage both health and SCNs may improve outcomes such as mortality and quality of life for people living with MLTCs (6, 7).

Care of people with MLTC is mostly managed in primary care where earlier studies show that 50% of consultations include social concerns. This is challenging given the rapid growth of people presenting with MTLCs. Stratified or group-based approaches, where people with similar health and SCNs are clustered together, offer a potentially efficient mechanism to identify those with the greatest needs and the highest risk of poor outcomes for prioritisation of interventions (8, 9). In the context of MLTCs, Artificial Intelligence (or AI) clustering has been used to improve clinical decision making. (10, 11). Our group recently derived clusters, using AI methodology, that are based on health and social care need. (7, 12) To impact on care delivery, these clusters need to be harnessed and implemented into a clinical intervention in primary care. This requires more detailed consideration as this will only be effective if informed by the views of potential users and stakeholders to maximise likely uptake, impact and adoption (13). In this study, we aimed to examine the acceptability and perceived value of AI-derived clusters by health and social care need in service users and their carer’s.

## Methods

### Study design, participants and setting

Two qualitative interview studies were conducted with 1) Individuals involved in the care of people living with MLTCs, including health and social care professionals (HSCPs) and unpaid carers, and 2) People living with MLTCs. Ethical approval was granted by the Research Integrity and Governance team and Faculty of Medicine Ethics Committee at the University of Southampton (87759).

Inclusion criteria:

- Aged 18 years or over
- English speaker
- UK resident
- **Either** a person living with more than one long-term health condition (self-identified by the patient as defined by our list of 59 conditions (14), developed with patient and stakeholder involvement); **or** people involved in the care of people living with MLTCs e.g. primary care clinicians (GPs, nurses, social prescribers), voluntary, charity sector and private providers, **or** unpaid carers of people with MLTCs. Participants were characterized by their primary motivation to join the study if they identified with multiple roles.

Exclusion criteria:

- Lacked capacity to provide informed consent.

## Data collection

Recruitment ran from September 2023 to March 2024. Data collection stopped when we had achieved theoretical saturation and no new concepts, codes or themes were apparent (15).

Participants were recruited via a community approach. This involved adverts (see Appendix 1) on social media platforms, university websites and newsletters, and established networks (with consent from gatekeepers) including city councils, adult social care organisations, local and national charities, university interest groups, libraries, food banks, GP education networks, Clinical Research Networks and care homes. Participants were invited to share the research within their own peer networks.

Individuals contacted the lead researcher (SZH) or accessed the study website directly to express interest, review information about the study and complete an online consent form and demographic questions. Eligible participants were contacted by the lead researcher to schedule the interview. Purposive sampling was used to ensure we captured experiences from people living with different combinations of MLTCs, staff working in different sectors of health and social care and across a range of demographics (e.g. sex, age, ethnicity).

A topic guide (Appendix 2) was developed based on our study aims, previous research, expertise within the team and piloting with the target populations prior to use. Terms and concepts were explained in lay terms as part of the interviews, including a description of AI-derived clusters. Interviews were carried out remotely (either online or by telephone) to enable geographical diversity. In-person interviews were offered as an option, however, no participants opted to participate in this way. Interviews were conducted by an experienced female qualitative researcher (SZH), and MSc level students trained in qualitative research. Interviews lasted between 21 and 102 minutes. Interviews were audio recorded, transcribed verbatim and anonymised. Each participant was reimbursed for their time with a £25 voucher. Data were transcribed by MSc level students and a third-party transcription service, adhering to GDPR (General Data Protection Regulation).

## Data analysis

Analysis was led by SZH and LM, with input from the wider team. Analysis began with a familiarisation process of reading and re-reading the interview transcripts. Data from interviews with people involved in the care of MLTCs were analysed first, using an open coding process via Reflexive Thematic analysis (16). The coding was facilitated by NVivo 14 and documented in an initial coding manual to support discussion with a stakeholder team. Preliminary themes were developed from the codes by identifying patterns of shared meaning, these were then applied to transcripts from people living with MLTCs using a codebook approach to thematic analysis. Themes were developed and refined to incorporate additional codes generated recognising that HSCPs, carers and people living with MLTCs have different experiences of MLTCs and SCNs.

Consensus on the thematic structure and richness of the data to address the study aim were confirmed through discussion with the wider stakeholder team. Researchers involved in analysis also felt that, at this stage, no new codes or themes were being developed, and that theoretical saturation was met. We remained open to new concepts, codes or themes throughout the analysis; no new insights were developed from the data during the latter phases.

## Public Involvement

Two individuals with lived experience of MLTCs provided input into the public-facing study materials, reimbursement arrangements, and overall study direction. This included changing the way we addressed “Individuals involved in the care of people living with MLTCs" in all study materials. Originally, this was worded as “managing” the care of people living with MLTCs. Our public contributor felt that this language created a power imbalance between health professionals and was disempowering for people with lived experiences. Our public contributors also helped to ensure that our interpretation of the data was grounded in the participants’ experiences and that the interpretation was able to have meaningful impact beyond the study.

## Results

### Participants

44 interviews were completed, with 24 people living with MLTCs and 20 people involved in the care of MLTCs. Table 1 outlines the characteristics of the people living with MLTCs and Table 2 outlines the characteristics of participants involved in the care of people living with MLTCs. People living with MLTCs reported living with an average of 4 total conditions (ranging from 2 to 11 conditions). The most common MLTCs were:

- Long-term musculoskeletal problems due to injury.
- Depression.
- Anxiety.
- Chronic primary pain.
- Post-Traumatic Stress Disorder.

**Table 1:** Demographics for people living with MLTCs (N=24)

|  | Demographics | N |
| --- | --- | --- |
| *Gender* | Woman | 18 |
|  | Man | 5 |
|  | Non-binary | 1 |
| *Age* | 18 to 29 | 3 |
|  | 30 to 39 | 5 |
|  | 40 to 49 | 4 |
|  | 50 to 59 | 9 |
|  | 60 to 69 | 1 |
|  | 70+ | 2 |
| *Ethnicity* | White – British | 14 |
|  | Asian or Asian British – Indian | 2 |
|  | White – Any other White background | 2 |
|  | Other ethnic groups – Any other ethnic group | 2 |
|  | Black or Black British – Caribbean | 1 |
|  | Asian or Asian British – Pakistani | 2 |
|  | Mixed – White & Asian | 1 |
| *Occupation* | Retired | 5 |
|  | Unemployed/Disabled and unable to work | 6 |
|  | Student | 4 |
|  | Volunteer | 2 |
|  | Full time employment | 7 |
| *Education* | GCSE | 1 |
|  | A Levels | 4 |
|  | Undergraduate | 8 |
|  | Postgraduate | 8 |
|  | Other qualification | 3 |

**Table 2:**
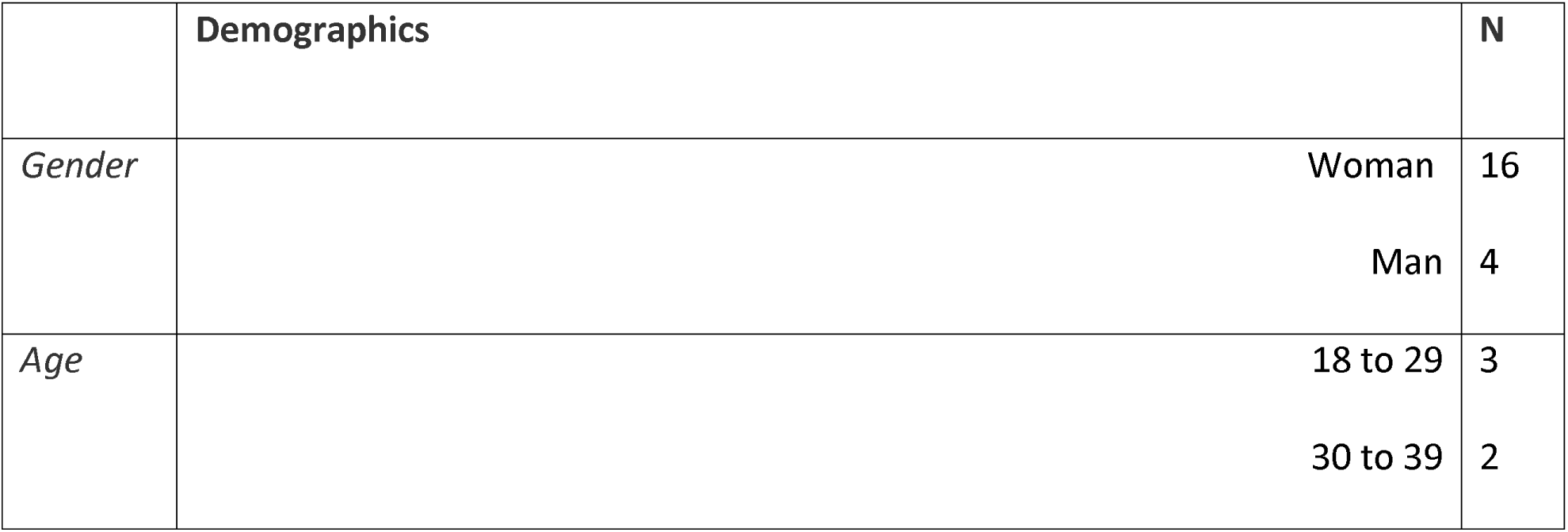
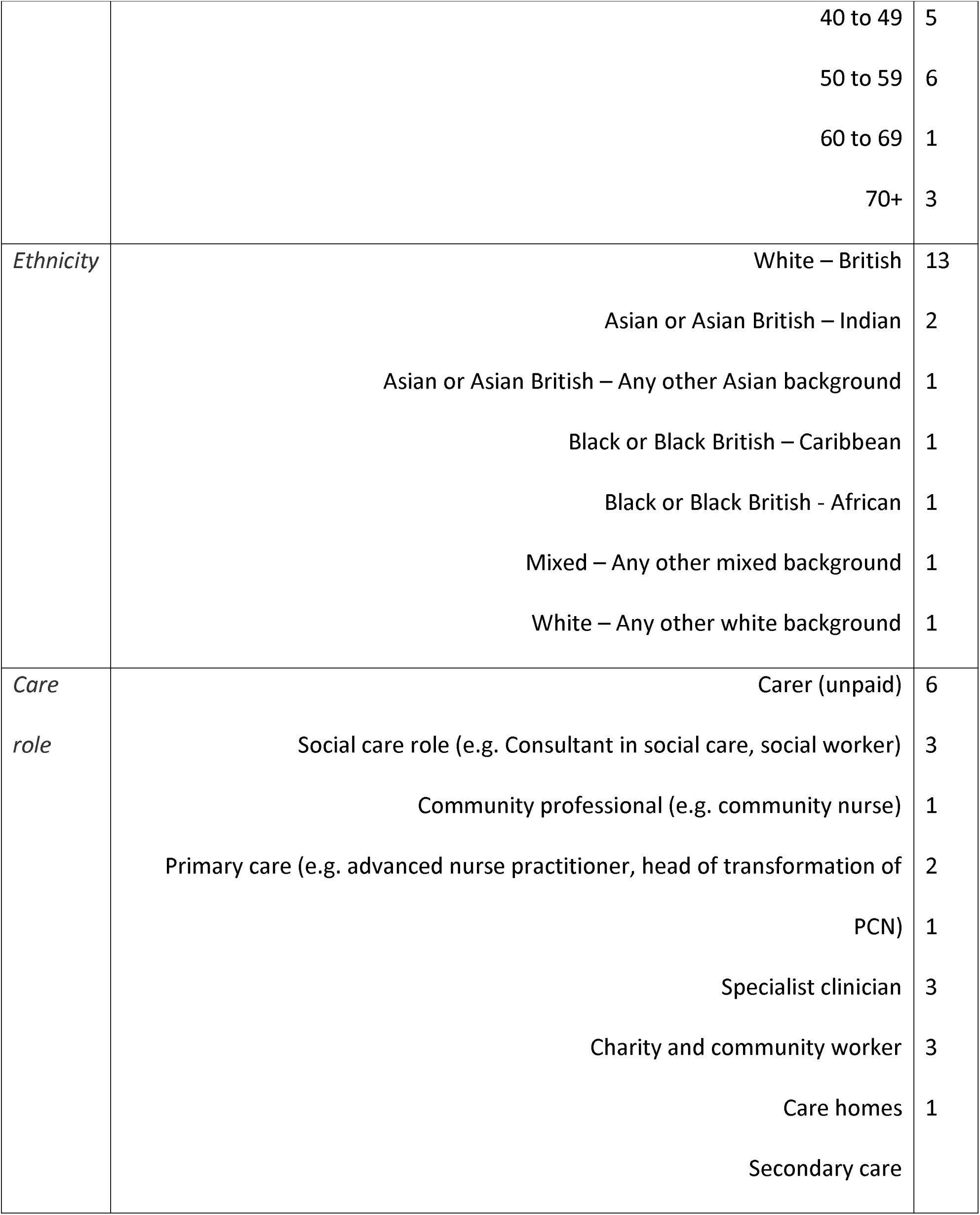
Demographics for people involved in the care of MLTCs (n=20)

Four themes were developed from the data. Primary care was commonly viewed as the starting point for discussion of SCNs (Theme 1), but the capacity for services to support SCNs was perceived as limited (Theme 2). AI was considered to be an efficient approach to deliver holistic care (Theme 3) when used to supplement effective, clinical conversations (Theme 4).

## Theme 1: Primary care is the ‘starting point’ for discussing social care needs

MLTCs are seen as complex; patients described struggling to understand which of their SCNs are ‘caused’ by which of their multiple conditions. This led to patients not knowing which speciality-specific clinician is best to speak to first when SCNs occur. However, when prompted, all participants saw primary care as the default starting point for these discussions, with GPs being seen as providing the best ‘general’ all-round knowledge. Furthermore, people involved in the care of MLTCs speculated that people living with MLTCs might feel reluctance towards accessing social care services when addressing their SCNs. HSCPs in particular, believe that people living with MLTCs perceive social care services to be only for people with ‘extreme’ SCNs.

> *Something about how having different conditions can make it harder to work out what’s going on* (Person living with MLTCs, 07)

> *Maybe a GP would have an important role there because a good GP would know their patient and would know what they struggle with and probably be able to speak to them.* (Person living with MLTCs, 17)

> *A lot of people don’t always come straight to social work when it comes to issues like this. In fact, if possible, they’ll try and give social work a wide berth and try and use other services instead.* (HSCP 09, student social worker and carer)

Despite participants viewing primary care as a starting point, barriers were identified which led to people living with MLTCs navigating their SCNs alone. Identified barriers included struggles getting GP appointments, and short appointments that prioritised health-related needs over SCNs.

> *How much time have they got for them? Fifteen minutes? It’s been extended now from 10 to 15 minutes, but they maintain that you go to them with one thing, don’t come with a list, but you can’t help that because with long-term conditions, one condition will affect another.* (Person living with MLTCs, 02)

> *Often people struggle to see the GP and get into appointments, so they will struggle with accessing things.* (HSCP 05, community nurse)

People living with MLTCs, HSCPs and carers felt that the expansion of roles within primary care could open the opportunity for more conversations about SCNs. The use of expanded roles in facilitating these conversations was identified by all participant groups without prompting, suggesting that they felt this was a viable strategy when considering how to encourage conversations about SCNs within primary care. Roles within primary care that participants felt could be used for these conversations included social prescribers or nurses. Participants felt that using these roles for conversations about SCNs could help to address the barriers currently experienced in accessing GP appointments.

> *We have social prescribers now. We have a whole range of healthcare workers who work now in primary care, in GP practices, and I think that is a good thing.* (HSCP 20, community involvement manager)

> *It’s debatable whether it should be the GP because they’re so busy that they don’t seem to have the time. It’s difficult to get appointments, etc. So, I wonder whether or not there should be a dedicated person in maybe each surgery.* (HSCP 02, primary care admin and carer)

> *I’m guessing a doctor wouldn’t be able to do it because they’re far too busy. Perhaps it would have to be a nurse.* (Person living with MLTCs, 15)

## Theme 2: Social care needs are rising, but the capacity in all NHS systems is decreasing

HSCPs expressed a desire to help patients address their SCNs but felt limited by their current capacity and lack of resources (e.g. time, available GPs). HSCPs report receiving a lack of training on SCNs and felt unsure about the types of support available to signpost towards. HSCPs also expressed feeling unaware of what help patients should expect to receive from each service and lacked confidence in knowing things like the financial support available. HSCPs felt that most signposting information related to SCNs is learned on the job, which may lead to inconsistent care between individuals with the same condition.

> *Going to primary care, I find the training is definitely reduced when compared to the hospital setting. You don’t know what you don’t know in terms of services that are out there for people or support that people can access.* (HSCP 06, primary care nurse)

> *Sometimes GPs don’t know what support is available, so if a computer were to tell the GP, ’Offer this person this support,’ then that could be helpful.* (Person living with MLTCs, 20)

All participants recognised that signposting information is frequently changing with new services being added and old services closing. HSCPs felt that, keeping information up-to-date and relevant is something they do not have the capacity to manage without adding to the overall burnout and fatigue they already experience. People living with MLTCs and their carers also felt that keeping up to date with SCNs resources within the constantly evolving social care landscape was not realistic for primary care professionals.

> *I think anything that you can do to, not make our jobs easier, but I guess it is, but also to make us more efficient because we’re just so - you go into work and literally you don’t stop from the moment you get in to the moment you leave. You’re always late, half the time you’re working through your lunch.* (HSCP 06, primary care nurse)

> *I think I like the idea that the GP would have that information on hand, but I think it’s a big ask of GPs to be up-to-date with all of it.* (Person living with MLTCs, 07)

## Theme 3: AI offers an efficient approach when used cautiously to deliver holistic care

When introduced to the idea of AI-based clustering, all participant groups felt that the approach held promise for improving system efficiency and getting patients support for SCNs. They felt the approach could help to identify SCNs earlier, prevent SCNs from developing into a more severe problem, help to tailor interventions to be more specific for patients, improve signposting efficiency and help with the allocation of funding and services based on patient need. There was a sense that, if AI can help to make this process quicker and more efficient, patients, carers and staff would be accepting of the NHS using it within the primary care context.

> *If we can cluster patients together and provide support that’s appropriate between us, it’s only going to benefit the patients.* (HSCP 05, community nurse)

> *It sounds like the good example of where technology can help to work. That would take a long time for staff to do, so and it also sounds like it might sort of be edging towards a more holistic view of patients rather than just the kind of silo based structure.* (Person living with MLTCs, 18)

Concerns about the use of AI within the NHS must be addressed to achieve the promise of improved efficiency. Participants’ main concern was that the data might not be 100% reliable and decisions about their care might be based on data that they would consider to be false. Participants felt that not all SCNs conversations within primary care are coded, and not all SCNs conversations take place within primary care. Therefore, the data representing SCNs might not be fully complete or representative of their experiences and engagement with SCNs conversations and services. All participant groups also felt that human input would be needed to ensure the AI is working correctly and in-line with clinical judgement.

> *I think that there could be mistakes made. That’s the problem, I suppose. I suppose, the algorithm would be quite reliable, but there’s always going to be something a bit different about people, so it might not pick that up.* (HSCP 02, primary care admin and carer)

> *The biggest problem is that the data that the NHS holds is very patchy. It has loads of holes in it and it’s not up to date, and it has less data about some people than other people… there’s a bit of a risk, isn’t there, that if you’re making those deductions or forming those groups or forming conclusions based on patchy information, you could end up with conclusions that haven’t been tested out or aren’t right.* (Person living with MLTCs, 07)

All participant groups felt that the use of AI was acceptable in primary care, as long as the data remained secure within the NHS and was not used by third-party companies. There were concerns about the data being used in a way that could discriminate or marginalize groups, and they were keen for processes to be put in place to mitigate against this.

> *I think the only thing I would have pause is if that data falls into the hands of, for example, insurance companies.* (HSCP 07, primary care)

> *A lot of people are very mistrustful of AI and the storage and the security of that, so I suppose there might be some mistrust and some backlash over the use of technology in that way.* (Person living with MLTCs, 04)

> *You don’t know who’s got access to the data; how it’s going to be used; what other things will it affect? Will it be something that just the GP has access to?* (Person living with MLTCs, 16)

## Theme 4: AI could be used to supplement effective, clinical conversations

Despite positive views on the potential of the AI-cluster approach, all participant groups felt that AI cannot account for all individual differences, such as the psychosocial and contextual background that each person brings to a consultation based on their previous experiences. There was a desire for care to feel meaningful for each individual, with patients feeling that they have their own unique experiences, values and needs, which should be addressed. There was a sense that grouping people could be an efficient way to structure care, but the needs of the individual should remain at the heart of the consultation. All participant groups highlighted the importance of having an effective conversation to accompany the AI tool. Patients stated that they value feeling listened to and validated when discussing their SCNs.

> *I think that often, people aren’t necessarily listened to about the support that they actually need. I think if we listened to the people that were struggling, it would be a lot better.* (Person living with MLTCs, 11)

> *Again it’s a big issue but in an ideal world people shouldn’t have to fight to get seen or to be listened to. Even if a GP has to sit there and say ’I can refer you but the waiting list is years long’ at least they’ve listened to you and taken on board that you need a referral.* (HSCP 18, care home staff)

> *I think that knowing that somebody cares and is listening to you, 1) is quite important and really helpful, 2) feeling that there is some hope*. (Person living with MLTCs, 17)

Some patients expressed struggles in discussing their future risks of developing SCNs. They felt this could be framed in a negative way, which may cause them to feel anxious about the future and helpless to address their increasing SCNs. All participant groups acknowledged that the best approach to discussing future risks would involve a conversation framed in a positive manner, whilst remaining open, honest and adaptive to the patient’s preferred communication style and needs.

> *I think ’risk’ could be quite worrying for the patient… people maybe get a bit of anxiety about that.* (HSCP 10 – physiotherapist)

> *I think the only downfall is the different mechanisms, different mediums of trying to communicate, trying to reach out, and trying to offer services to people. How useful is one resource going to be over the other? You have to consider the audience as well.* (Person living with MLTCs, 03)

## Discussion

In this study, we explored the acceptability and perceived value of an AI-derived clustering approach to identify health and SCNs in primary care. We interviewed patients, carers and health and social care professionals. The rationale was to inform intervention development that sufficiently considers individual health concerns and/or behaviours in supporting MLTC care (17). In our study, primary care was an acceptable context for conversations about SCNs, with GPs, nurses and social prescribers identified as having a potential role. There was a lack of training provided in primary care about SCNs and challenges with navigating the constantly changing and evolving services available for signposting. Systematic review evidence about how GPs manage patients with MLTCs supports these findings, with practitioners struggling with the fragmentation of services and clinical uncertainty when applying single-condition guidelines (18). There is an identified need for tools to support these processes within primary care.

Our study found that an AI approach was considered acceptable to identify patients where conversations about SCNs are required. However, concerns about data reliability and security were highlighted, which has been reflected in other qualitative research about the use of AI to structure healthcare (19–21). Patients in our study also strongly emphasised their desire to feel ‘listened to’ within primary care consultations, with a fear that AI-derived clusters may undermine a person-centered consultation. In a recent mixed-methods study, Witkowski et al. also noted a fear of ‘losing the human touch’ when using AI-based tools, conflicting with patients desire for person-centred care (22). Therefore, we suggest that AI cannot achieve the level of personalization desired by patients when used alone and needs to be used in combination with effective clinical conversations. These need to be supported by behavioral and psychological evidence to promote engagement. This is further supported by a recent literature review, citing the importance of the ‘assistive use’ of AI in healthcare (23).

Overall, our findings suggest a complimentary approach that may involve AI-derived clusters combined with a ‘human’ conversation tool to facilitate effective SCNs discussions within primary care. These findings are important to aid our understanding about the current challenges of discussing SCNs and thoughts on AI-cluster approaches to develop interventions that are acceptable, feasible and perceived as valuable by service users.

## Key recommendations

These findings suggest that there are various factors to consider when discussing SCNs within primary care, but people living with MLTCs and people involved in the care of MLTCs perceive these conversations to be of value. This indicates a desire to improve care for SCNs for people living with MLTCs, but a lack of effective ways to currently facilitate this within the current capacities of primary care. Based on the findings, we have several key recommendations for considering the use of AI-cluster interventions within the primary care context to improve the identification of SCNs. AI interventions should:

- Be brief to avoid adding to the existing workload of primary care staff.
- Be adaptable, to be used by different staff members within primary care.
- Contain information that is quick and easy for clinicians and patients’ to use, and is kept up to date.
- Be used as a tool to assist in identifying risk in primary care, but should be used in combination with effective conversation strategies or interventions to ensure patient care remains personalised.
- Support primary care professionals to communicate SCNs risk to patients in an engaging way, where the focus is around empowerment of individual choice.
- Provide support for managing reluctance to have conversations about SCNs from both the clinician and patient perspective.

## Strengths and limitations

To our knowledge, this is the first study to explore patient, carer and HSCPs views on AI-derived clustering approaches to address SCNs. We used qualitative data from patients, carers and HSCPs to provide a rich understanding alongside including diversity within the patient group and representation of the sample to capture disparate views. Participants were, however, limited to proficient English speakers due to budget constraints limiting the availability of translation services. Our sample may not therefore fully reflect the full scope of primary care users across the country. Future research should, where possible, include views from participants who are proficient in languages other than English. Furthermore, considerations need to be given to individuals who are most deprived and in need, who may have incompleteness of data and recording Index of Multiple Deprivation (IMD). Careful consideration is needed to ensure the use of AI-derived clusters does not exacerbate inequality of access. Future research should consider using alternative recruitment strategies to capture the views of underserved populations to truly reflect the full scope of SCNs.

Only a limited number of primary care professionals, were recruited for this study, and no GPs were able to be included. This occurred despite an adaptive recruitment process and attempts to improve our approach. Therefore, the findings might not fully capture their views and experiences. However, our stakeholder group and core research team included several academic GPs, who were involved in the interpretation of our results.

The study topic, focusing on AI, may have limited interest in participation to only digitally literature people. Therefore, the views of people such as elderly people or people with specific health and cognitive needs, might be lacking from the data. This may have excluded views from people with the greatest social care need and should be consider in future recruitment strategies.

## Conclusion

An intervention based on AI-derived clustering to help aid discussions about SCNs within the primary care context is seen as potentially valuable by patients, carers and health care professionals, but there were some concerns around data security, completeness and ensuring care remains personalised. AI interventions could be used as an additive tool to improve the identification of patients at risk of developing SCNs. However, AI should always be supported by effective listening and tools to enable patient-centered conversations. Future interventions should also consider how AI-cluster approaches can be used to support and develop the capacity of multi-professional working when considering SCNs for patients’ living with MLTCs.

## Sources of financial or material support for the work

### Funding

This study is independent research funded by the National Institute for Health Research - the Artificial Intelligence for Multiple Long-Term Conditions, or "AIM". ’The development and validation of population clusters for integrating health and social care: A mixed-methods study on multiple long-term conditions’ (NIHR202637). The views expressed in this publication are those of the authors and not necessarily those of the NHS, the National Institute for Health Research or the Department of Health and Social Care.

This study/project is funded by the National Institute for Health and Care Research (NIHR) School for Primary Care Research (project reference 667 FR6). The views expressed are those of the author(s) and not necessarily those of the NIHR or the Department of Health and Social Care.

AF is supported by the National Institute of Health Research (NIHR) Oxford Biomedical Research Centre (BRC).

## Data Availability

All data produced in the present study are available upon reasonable request to the authors.

## List of abbreviations

SCNs: social care needs
HSCPs: health and social care professionals
NHS: National Health Service
NIHR: National Institute of Health Research
GP: General Practitioner

## Acknowledgements

We acknowledge the contribution made by stakeholders and PPI members (Firoza Davies and Adrian Richardson) who were involved in this work and we thank them for their input and support.

## Appendix 1 – Study adverts

Note:

This document outlines example text and/or images that we will use in our study advertisements. This may take the form of text, images and/or video content (with voiceovers). The adverts may evolve over time to match the developing needs of the study. However, the underlying meaning of the content will remain the same. These materials will include relevant ERGO numbers, dates and version numbers where possible (e.g. on images, in videos). Where wordcount is limited (e.g. on social media), participants will be linked to the study website where all ERGO numbers, dates and version numbers will be visible.

### People living with MLTCs

#### Advertisement for websites/posters

Do you have two or more health conditions? If yes, we are interested in hearing your views about a new approach to improving health, lifestyle and social care!

We’d like to chat with you on the telephone or online for up to 60 minutes. This will on a convenient date and time for you. We might be able to speak to you in-person, if you’d prefer.

If you are interested or have any questions, please email us at: [study email here] or phone: [study phone number here]

Or go to our website: [study website link]

#### Advertisement for social media (280 character word limit)

Do you have two or more health conditions?

We are interested in hearing your views about a new approach to improve care.

We’d like to chat with you on the telephone or online.

Email: XXXXXX@XXXXX Phone: XXXXXXXXXX

Or visit: [study website link]

### Health and social care professionals

#### Advertisement for websites/posters

Are you a health or social care professional who is involved in the healthcare of people with multiple long-term conditions? This includes doctors, nurses but also people who care for someone with multiple long-term conditions (either paid or family carers).

If yes, we are interested in hearing your views about a new approach to improving care by clustering people together who have similar health and social needs.

We’d like to invite you to participate in an interview either on Microsoft Teams or by telephone. We will give you some options of dates and times to make this convenient for you. This interview will last up to 60 minutes.

If you are interested or have any questions, please email us at: [study email here] or phone: [study phone number here]

#### Advertisement for social media

Are you a health or social care professional who is involved in the healthcare of people with long term conditions?

We’d like to hear your views on a new approach to improving care by inviting you to an online or telephone interview.

Email: XXXXXX@XXXXX Phone: XXXXXXXXXX

Or visit: [study website link]

## Appendix 2 – Topic Guide

### People living with MLTCs

#### Lay title: Exploring a new approach to improving health, lifestyle and social care

##### Introduction

- Explain who you are [your name, role & that you are part of a team at UoS].
- We would like to understand your experience living with several different long-term health conditions. We’d also like to know about how your health and lifestyle needs are (or are not) currently being met and why this might be happening. Then we’d like to chat to you about a new approach to support patients getting the help they need for their illnesses, and what your thoughts are on this approach.
- We will use the information from our talk to see if and how care can be improved for people with multiple health conditions in the UK.
- Thank you for completing the online consent form. Did you have any questions?
- As mentioned in our consent form, we will record our conversation so we can listen again to what is being said. Everything we talk about here will be **confidential**. We will take care to make sure that all the information you share with us is kept safely and securely. Your care providers will not know you have spoken with us.
- When I start the audio recorder, I will begin by confirming that you read and completed the online consent form and that you are happy to speak with me today. This is just so we have a verbal record of your consent, as well as that written version you completed online.
- I will take some notes during our interview, but the recording allows us to know exactly what you’ve said in your own words. We will remove your name and personal details so people will not be able to identify you.
- You may see me look over to one side during the interview, this is just because I am checking my questions on my other screen.
- The interview will last up to 60 minutes [check that the interviewee has this time available].
- The interview can be stopped at any time and without reason [if this happens, make a note of why they stopped it, if they mention].
- Before we start, may I please ask whether you are feeling comfortable and have everything you need, such as drinks or snacks, and comfortable room to talk.
- [start audio recorder] Now that I have the audio recorder on, can I confirm that you completed the online consent form for the study and you are happy to take part?

| Main Questions | Prompts Questions | Ideas that might be raised by the participant |
| --- | --- | --- |
|  |  | These will not be used as prompts/probes but will be for researchers to keep in mind if any of these topics are raised by the participant |
| **General context**<br>Please tell me about what it is like living with different long-term health conditions | • What conditions do you have?<br>• Talk me through anything that stops you from doing what you would like to do day-to-day, related to your health conditions<br>• Tell me about if you've ever sought any support for these issues.<br>• Tell me about any services or support that you think would improve your health conditions or your day-to-day life | Warm up question but move onto next section<br>ASAP – revisiting any ideas that emerge during the general context conversation |
| **Cluster approach**<br>We are looking for | • What do you think of this idea? | Prompt around the seven social care needs categories: |
| ways to help people think about their day-to-day needs when managing their health conditions.<br>To do this, routine data in the NHS (such as GP records) would be used to look for patterns to see which people have similar needs or challenges in their day-to-day life, to work out what other support they may need and when they might need them.<br>These patterns are going to be created | • How would you feel about being put into a group?<br>• Based on the information you gave on your online survey, our AI would place you in a group as being at risk of developing needs related to [x]. What do you think about that?<br>Does that reflect your experience of having these conditions? What do you think of the word 'risk'?<br>• What would you think if you were given advice about X social care need? Is that important to you? Why/why not?<br>• How would you feel if you didn't agree with the group you've been put | • ADL<br>• Community<br>• Disability<br>• Finance<br>• Mobility<br>• Residential<br>• Social Network |
| using artificial intelligence (or high tech computers).<br>You might have a similar pattern to multiple other people in the data.<br>We call this being in a 'group'. This isn't a group that ever meets, like a social support group. It just means that you have a similar pattern of data and needs to a group of other people.<br>By looking at the patterns in these groups, the NHS can see if people have | into? (give examples e.g. mental health support if they have self-identified low mental health needs)<br>• What are the problems with this approach? How could we address this?<br>• What are the advantages of this approach? What would encourage you to use this approach?<br>• Would you like to receive information about social care services? Why?<br>• How would you like this information to be given to you? What are your priorities for receiving this information? What services would you like to see? What do you find |  |
| similarities in their illness over time, and use this to predict what might happen to other people within that group over time.<br>The idea of looking at these patterns/ or groups like this, is to provide the right information for different groups more easily, more quickly and make sure people with needs aren't missed.<br>This grouping system isn't currently used in the NHS. | difficult about that at the moment?<br>• How and what would you like to be told by your GP?<br>• What do you think of a leaflet/website?<br>• Who should be having these conversations with you? Where would be the best place for support? |  |
| These patterns can be used to tailor support for each group. This could take various formats (leaflet, website, conversations with a health care provider) |  |  |
| Vignette/example (note: there will be several different vignettes to help to prompt discussion with participants)* | • What do you think of that story? How well can you relate to any of it yourself? Do you disagree with any of it? Why?<br>• Have you thought about your own social needs before? Why? And what are they?<br>• Do you feel like you've |  |
|  | been able to access the services you need?<br>• Do you think you need more support? What support? Why/what do you find difficult about that at the moment? |  |
| Is there anything else that you would like to say that we haven't discussed so far in this interview? | *Possibly follow up on an interesting point(s) raised by the interviewee, which may have come into your mind or you think is worth revisiting* |  |
| Do you have any questions for us? | • *Do you wish to receive results of the study?*<br>• *Ask the participant if they can **recommend anyone else** who we could interview?*<br>• *Thank participant. Ensure they have **study contact*** |  |
|  | *information for any future enquiries. (Note: prepared some contacts if participants are upset by the interview, such as GP, charity helpline, etc).*<br>• *Voucher? Get address for posting.*<br><u>Close the interview.</u> |  |

*Vignettes

The purpose of the vignettes is to be able to prompt around the complex idea of cluster-based interventions for improving health and social care in a relatable way for study participants. Vignettes will develop as the clusters are defined (in a separate study within our research team) and as interviews are completed, to address the research questions.

An example vignette is presented here. Where gaps are indicated, demographics similar to the participant will be used. For example, common names used in ethnic groups, similar age range to participant, same gender identity/pronouns as participant etc. This is to ensure that the story presented in the vignette is as relatable as possible.

The vignette has three sections. The first is a general introduction and a usual care scenario. The second is introducing the idea of clusters. The third introduces potential ideas about how clusters might be used in practice to improve health and social care. After each section of the vignette, the research will stop to probe and ask questions.

Example vignette

| Section | Vignette | Probes |
| --- | --- | --- |
| Section 1<br>General | [name] is a [age] who has [MLTC similar to study participant]. | What do you think of that story? How well can you relate to any |
| introduction and usual care | *They have been experiencing some challenges. [use challenges similar to those mentioned by the participant in the interview]*<br>*• With walking to and from the shops*<br>*• With affording to pay the bills due to being on sickness leave from work*<br>*They want help with these challenges but don't know where to start. They decide to ask their GP about this. However, the GP says they will have to discuss these challenges next time, as the appointment has already been taken up by discussing changes needed to medication.* | of it yourself? Do you disagree with any of it? Why?<br>• Have you thought about your own social needs before? Why? And what are they?<br>• Have you had conversations like this with your GP before? Why/why not? How did these conversations go?<br>• Do you feel like you've been able to access the services you need?<br>• Do you think you need more support? What support? Why?<br>• What did you think of |
|  |  | how the GP handled this situation in the story? |
| Section 2<br>Cluster<br>introduction | Here's how the story could have been a bit different.<br>At the appointment, the GP asks [name] some questions about their life and how they have been getting on. Based on the answers to these questions, the GP says that [name] fits into a group of people who have similar challenges with their long-term conditions that put them at a risk of their day-to-day life becoming more challenging over time.<br>By looking at what has helped people similar to [name] in the past, the GP might be able to work out what could be helpful. | • What do you think of this idea?<br>• How would you feel about being put into a group?<br>• Based on the information you gave on your online survey, our AI would place you in a group as being at risk of developing needs related to [x]. What do you think about that? Does that reflect your experience of having |
|  |  | these conditions?<br>What do you think of the word 'risk'?<br>• What would you think if you were given advice about X social care need? Is that important to you? Why/why not?<br>• What are the problems with this approach? How could we address this?<br>• What are the advantages of this approach? What would encourage you to use this approach?<br>• What would you expect the GP to do next? |
|  |  | • Would you like to receive information about social care services? Why?<br>• How would you like this information to be given to you? What are your priorities for receiving this information? What information would you like? What do you find difficult about that at the moment?<br>• How and what would you like to be told by your GP? |
| Section 3<br>Ideas | The GP provides a leaflet with resources that other people in the same group have found helpful. | • What do you think of a leaflet? Is this what you expected? |
| about<br>how<br>clusters<br>might be<br>used in<br>practice | These resources include social support networks that [name] can get in touch with. | Why/why not?<br>• What services would you expect to see in this leaflet? What do you find difficult about that at the moment?<br>• What are the problems with this approach? How could we address this?<br>• What are the advantages of this approach? What would encourage you to use this approach?<br>• What would you expect the GP to do next? |

